# The effect of ambient temperature on worldwide COVID-19 cases and deaths – an epidemiological study

**DOI:** 10.1101/2020.05.15.20102798

**Authors:** Anver Sethwala, Mohamed Akbarally, Nathan Better, Jeffrey Lefkovits, Leeanne Grigg, Huzefa Akbarally

## Abstract

**Background:** The role of ambient temperature in the spread of SARS-CoV-2 infections and subsequent deaths due to COVID-19 remains contentious. Coronaviruses such as the 2003 SARS-CoV showed an increased risk of transmission during cooler days. We sought to analyse the effects of ambient temperature on SARS-COV-2 transmission and deaths related to the virus.

**Methods:** The world population of COVID-19 cases and attributable deaths from the 23^rd^ January 2020 to 11^th^ April 2020 were analysed. Temperature 5 days before cases and 23 days prior to deaths (to account for the time lag of incubation period and time from symptoms to death) was compared to the average temperature experienced by the world population.

**Results:** The total number of cases during this period was 1,605,788 and total number of deaths was 103,471. The median temperature at the time of COVID-19 infection was 9.12°C (10–90^th^ percentile 4.29–17.97°C) whilst the median temperature of the world population for the same period was 9.61°C warmer at 18.73°C (10–90^th^ percentile 4.09-28.49°C) with a notional p-value =5.1 x10^−11^. The median temperature at the time of a COVID-19 death was 9.72°C (10–90^th^ percentile 5.39–14.11°C) whilst the median temperature of the world population was 7.55°C warmer at 17.27°C (10–90^th^ percentile 2.57°C-27.76°C) with a notional p-value = 1.1 x10^−10^. 80% of all COVID-19 related cases and deaths occurred between 4.29°C and 17.97°C.

**Conclusion:** A definitive association between infection rate and death from COVID-19 and ambient temperature exists, with the highest risk occurring around 9°C. Governments should maintain vigilance with containment strategies when the ambient temperatures correspond to this highest risk.

## Introduction

SARS-CoV-2 is the causative pathogen for COVID-19 and was first identified in December 2019 after a series of cases of pneumonia were linked to a live animal and seafood market in Wuhan, China.^1^ It soon spread rampantly throughout the world resulting in a pandemic that has infected over 1.7 million people and led to over 100,000 deaths as of 12^th^ April 2020.^2^ While South East Asia may have been expected to be the region most affected due to its geographical proximity and travel relationship with Wuhan, it has been cooler, further regions of the globe such as Tehran, Milan and Madrid in February and New York and London in March that have experienced the highest burden of disease. Conversely, warmer regions in South East Asia and Africa have had the lowest outbreaks.^2^ This has raised questions about the effect of environmental factors such as temperature, humidity and ultraviolet radiation on the viability and rate of transmission of SARS-CoV-2.

Experiments on coronaviruses in controlled environments show that conditions conducive to its survival are at temperatures around 4°C and relative humidity between 20–80%.^3^ In these conditions, coronaviruses can survive up to 28 days. Inactivation of the viruses occurs rapidly above 20°C. When temperatures reach close to 40°C, the coronaviruses last only a few hours at most.^3,4^ Epidemiological data on environmental factors in the 2003 Severe Acute Respiratory Syndrome (SARS) epidemic^5–7^ showed a significant increase of up to 18-fold in case numbers on cooler days, suggesting that coronaviruses transmit more efficiently during colder weather.^6^

Although a number of studies have assessed the relationship between temperature and COVID-19 outbreaks, most were conducted early into the outbreak with low case numbers^8^, are pre-prints and not peer reviewed. We therefore sought to collate worldwide data to analyse the correlation of ambient temperature with COVID-19 cases and deaths. In order to analyse this relationship in detail, we also benchmarked the temperatures that the world population experiences.

## Methods

### Cases and deaths

We considered all cases and deaths related to COVID-19 from the 23^rd^ of January 2020 to the 11^th^ April 2020. Data for case numbers and deaths in each country/region were obtained from the COVID-19 Data Repository at Johns Hopkins Center for Systems Science and Engineering. The total number of cases during this period was 1,605,788, which represents the total population of cases and total number of deaths was 103,471, which represents the total world population of deaths.

### Temperature data

Historical temperature data were obtained from the Dark Sky weather database (powered by Dark Sky, https://darksky.net/poweredby/). In order to work out average daily temperature, we recorded hourly temperatures for each day in each country/region and computed the mean. This will be referred to as the average temperature. The United States of America (USA), China, Canada and Australia were subdivided into regions as per the International organization for standardization (ISO) 3166–2 standard and recorded temperatures at each of their respective regional capitals. This was done to facilitate the alignment of temperature data with data available from these countries on confirmed cases and deaths in each of their respective subdivisions. With India, there were no individual case numbers and deaths available by region. As it is a country that is comprised of many highly populated cities with diverse weather conditions, we separately subdivided India (according to the ISO 3166–2 standard) when determining the temperature distribution of the world population. For all other countries, we recorded the temperature of its capital city.

### Statistical analysis

Temperatures 5 days before cases (17^th^ January 2020 to 6^th^ April 2020) were taken to account for the time lag resulting from the median incubation period of SARS-CoV-2.^9^ Temperatures 23 days prior to deaths (31^th^ December 2019 to 19^th^ March 2020) were obtained to account for the mean duration from symptoms to death of 18 days^10^ plus 5 days for the incubation period of the virus.^9^ The results were collated using Python Software® and graphing done using Matplotlib (J. D. Hunter, “Matplotlib: A 2D Graphics Environment”, Computing in Science & Engineering, vol. 9, no. 3, pp. 90–95, 2007).

To compare the world population temperature data, we used the world population of 7.163 billion divided into countries/regions.^11^ The average daily temperatures in those countries from 17^th^ January 2020 to 6^th^ April 2020 for cases and 31^th^ December 2019 to 19^th^ March 2020 for deaths were obtained. We then scaled these data so that it totalled the same as the world number of cases and deaths respectively. This allowed us to plot these data alongside the cases and deaths graphs for comparison. A notional p-value was calculated by the Wilcoxon test, as the distribution of the world population and cases are both non-parametric distributions.

## Results

### Cases

Figure 1 shows that the distribution of average temperature at the time of infection for a confirmed case of COVID-19 was different to the average temperature distribution of the world population during the same time period. The median temperature for a COVID-19 case was 9.12°C (10–90^th^ percentile 4.29–17.97°C) while the median temperature of the world population for the same period was 18.73°C (10–90^th^ percentile 4.09–28.49°C) with a notional p-value = 5.1 x10^−11^. This is void of sampling error as the world population is 7.163 billion and the world population of cases is 1,605,788. The 9.61°C difference in temperature between the two samples represented a highly significant difference in temperature distributions between COVID-19 cases and the world population.

**Figure 1.**
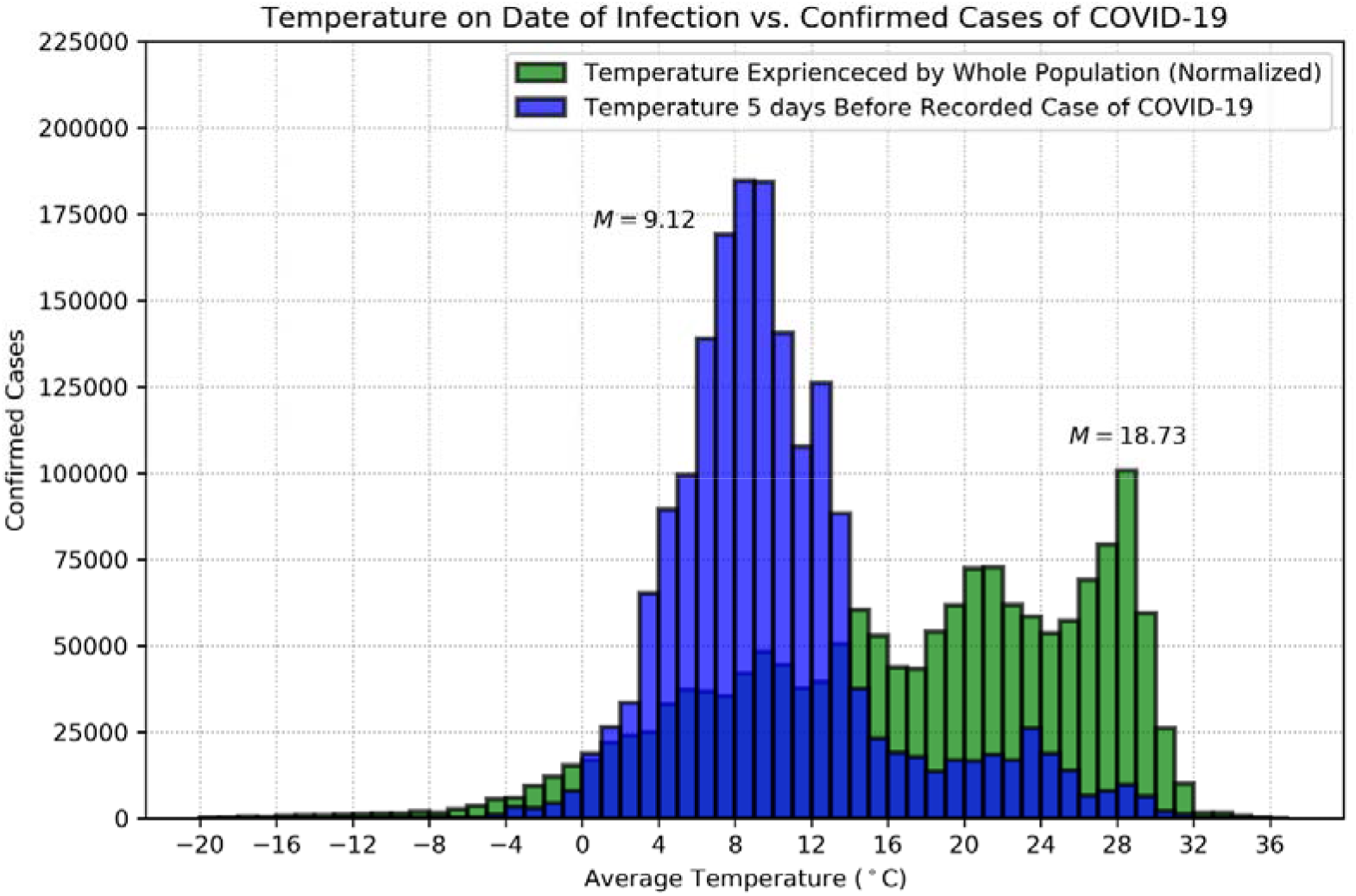
Comparison of temperatures on day of infection for global COVID-19 cases and world population (normalized). The graph shows that the median temperature experienced by confirmed COVID-19 case is 9.12°C compared to 18.73°C for the world population (notional p-value = 5.1 x10^−11^).

### Deaths

Figure 2 demonstrates the difference between the average temperatures at the time of infection for patients that died of COVID-19 compared with the average temperature distribution of the world population during the same time period. The median temperature for a COVID-19 death was 9.72°C (10–90^th^ percentile 5.39–14.11°C) similar to the median temperature for a confirmed case (9.12°C). In contrast, the median temperature of the world population for the same period was 17.27°C, (10–90^th^ percentile 2.57–27.76°C) which is 7.55°C warmer with a notional p-value = 1.1 x10^−10^. This is void of which is sampling error as the world population is 7.163 billion and the world population of deaths is 103,471. There was a strong relationship between average temperature and transmission rates of COVID-19, with at least 80% of the cases and deaths occurring between 4.29°C and 17.97°C.

**Figure 2.**
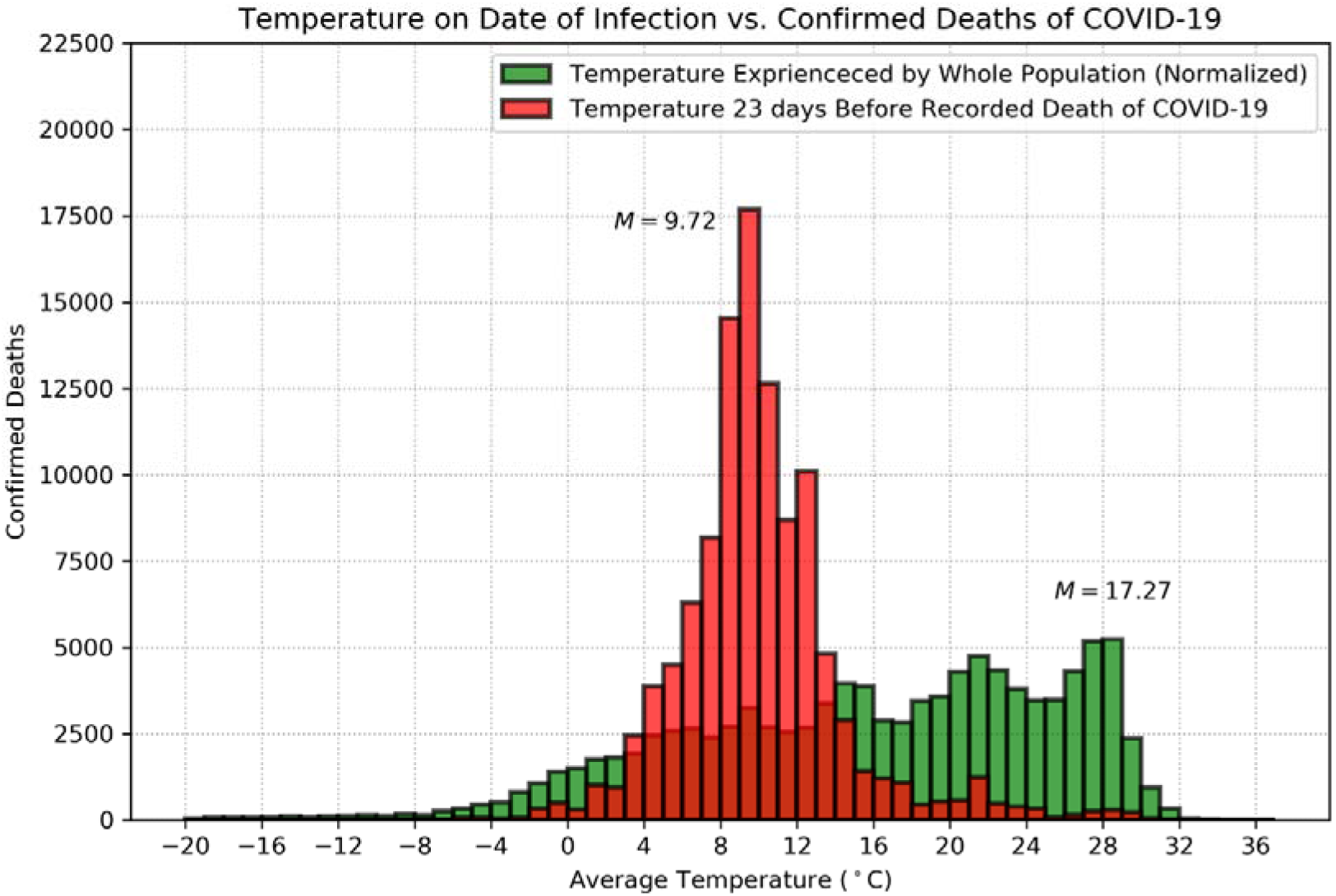
Comparison of temperatures on day of infection for global COVID-19 deaths and world population (normalized). The graph shows that the median temperature experienced by confirmed COVID-19 death is 9.72°C compared to 17.27°C for the world population (notional p-value = 1.1 x10^−10^).

Based on our results regarding ambient temperatures and cases and deaths, we explored the relationship of increased SAR-CoV-2 transmission as temperatures approached the median temperature of 9.12°C. Figure 3 demonstrates the world map with countries/regions shaded in the average temperature bands of the 80% CI (4.29–17.97°C), 60% CI (5.93–13.02°C) and 40% CI (7.22–11.50°C) around the median temperature of 9.12°C for worldwide cases during two separate weeks in our study period – Week 4 from 12/02/2020 to 19/02/2020 and Week 9 from 18/03/2020 to 25/03/2020. Superimposed on the map are black dots, each representing 400 new cases in the country / region. These bands were derived from the analysis of the world population of cases data presented in Figure 1. During Week 4, the black dots are localized mainly to China where most provinces are within the 40% CI band. During Week 9, the black dots are mainly in Europe and New York which are mostly within the 40% CI band. In order to study the temperature-outbreak relationship further, we selected 6 countries/regions {Hubei, Italy, New York, Singapore, Victoria (Australia) and Nigeria} from the world map and obtained the average temperature every 4 days from the 1^st^ of January 2019 to the 13^th^ of April 2020 (Figure 4). These 6 locations were selected after review of geographical data to reflect 3 locations with a high (Hubei, Italy, New York) and 3 locations with a relatively low {Singapore, Victoria (Australia) and Nigeria} prevalence of disease. When considering the 60% confidence interval band, Hubei province, China, was in this band from mid-January until mid-February 2020 – the period in which there was an uncontrolled increase in cases seen in Wuhan, China (Figure 5). Italy was in the same temperature band from mid-November 2019 to end-March 2020. As infection cases escaped China and reached Milan, Italy, temperature conditions were ideal for early uncontrolled growth, eventually occurring in late February and early March (Figure 5). New York, another city with large numbers of international travellers, was significantly colder than Milan in early February and only reached temperatures favourable for virus transmission at the end of February and maintained them throughout March. Corresponding to this temperature pattern, New York experienced its phase of uncontrolled growth in cases in late March/early April (Figure 5). So far, Singapore, Victoria (Australia) and Nigeria have not entered the at-risk temperature band (Figure 5).

**Figure 3.**
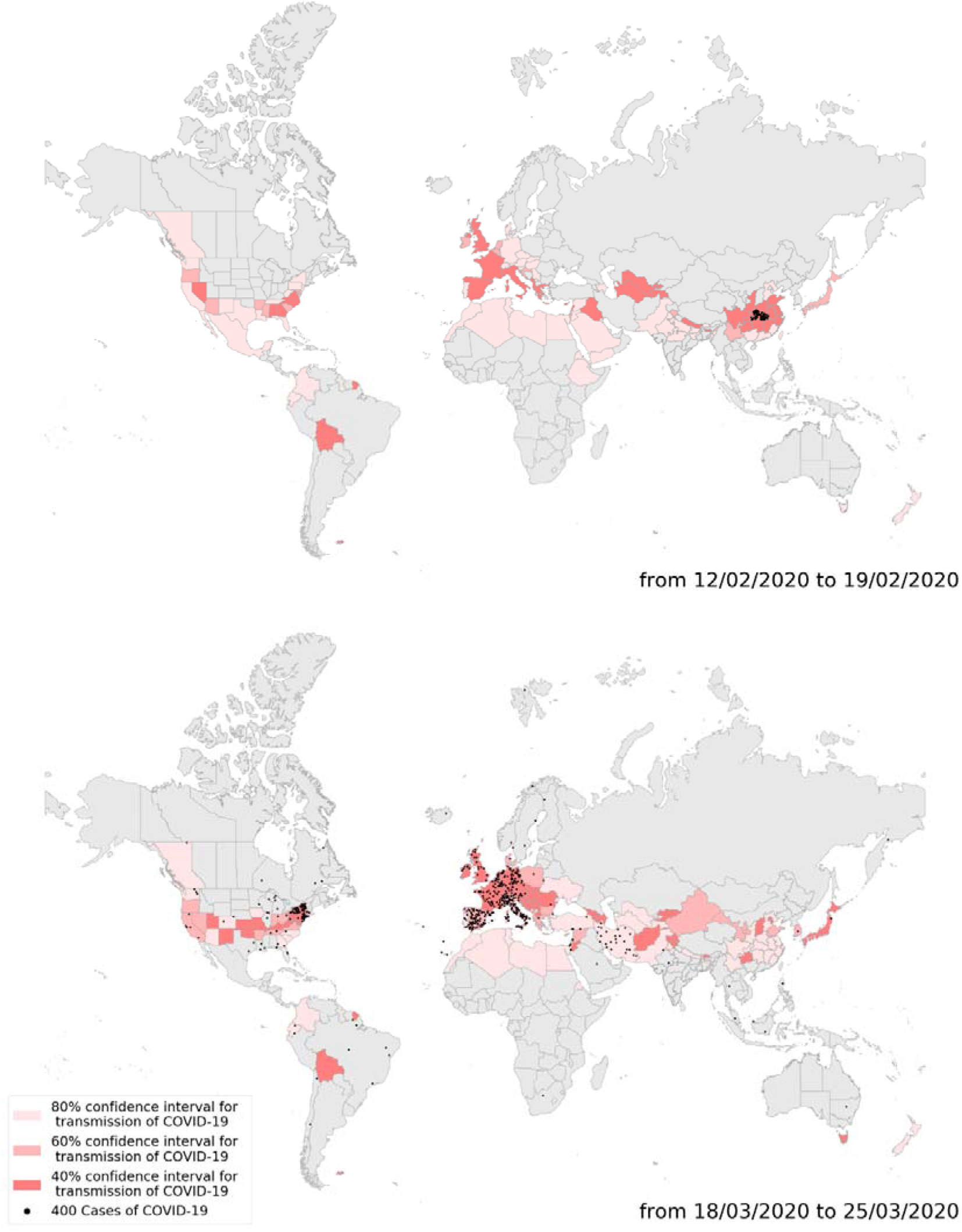
World map showing temperature bands around the median temperature of 9.12°C superimposed by new cases of COVID-19 (black dots) during two separate weeks in the outbreak. Temperatures 5 days before cases are used to account for the incubation period of SARS-CoV-2.

**Figure 4.**
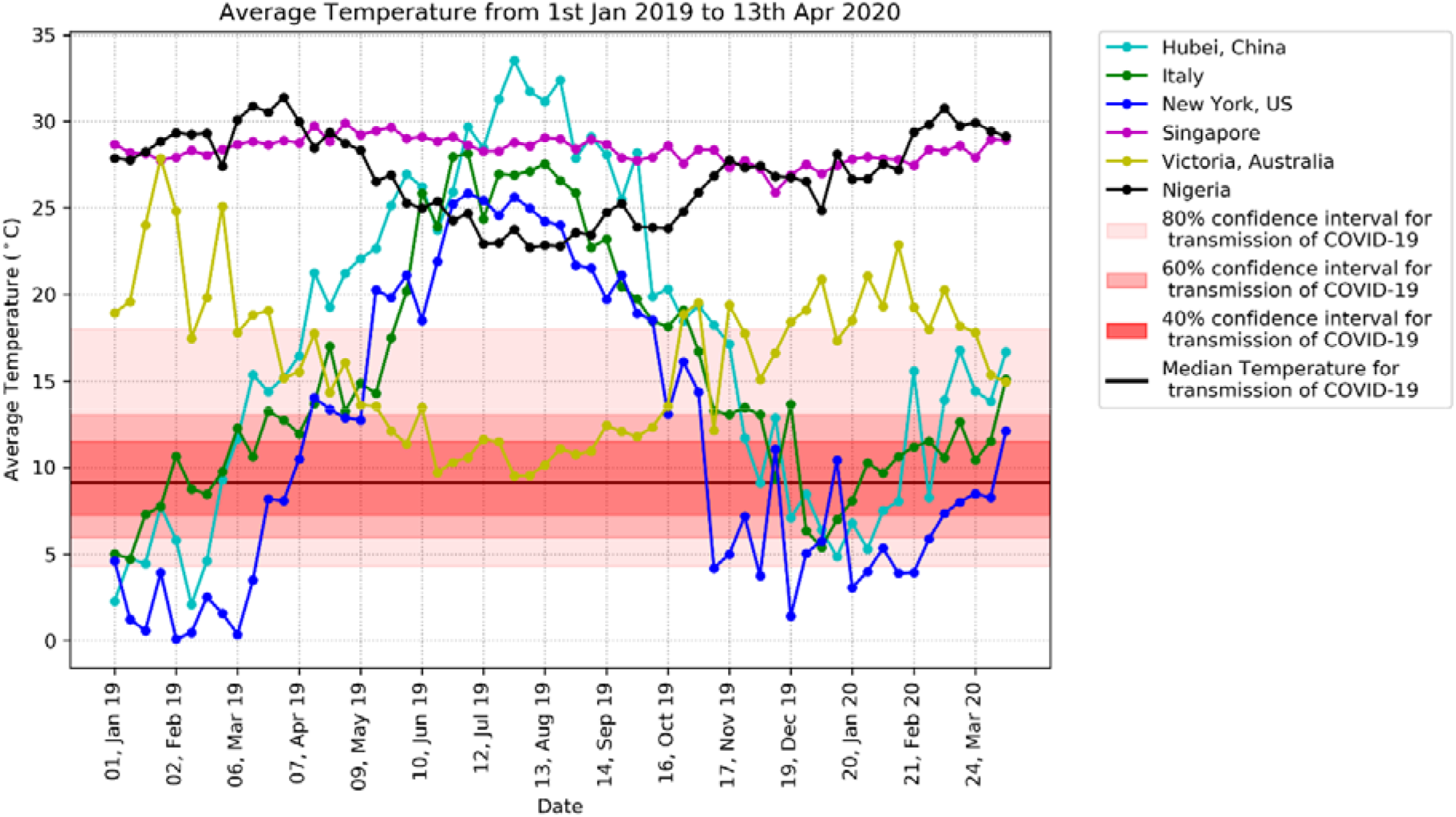
Temperature trends in particular regions with time showing their relationship around the median temperature of 9.12°C for cases of COVID-19

**Figure 5.**
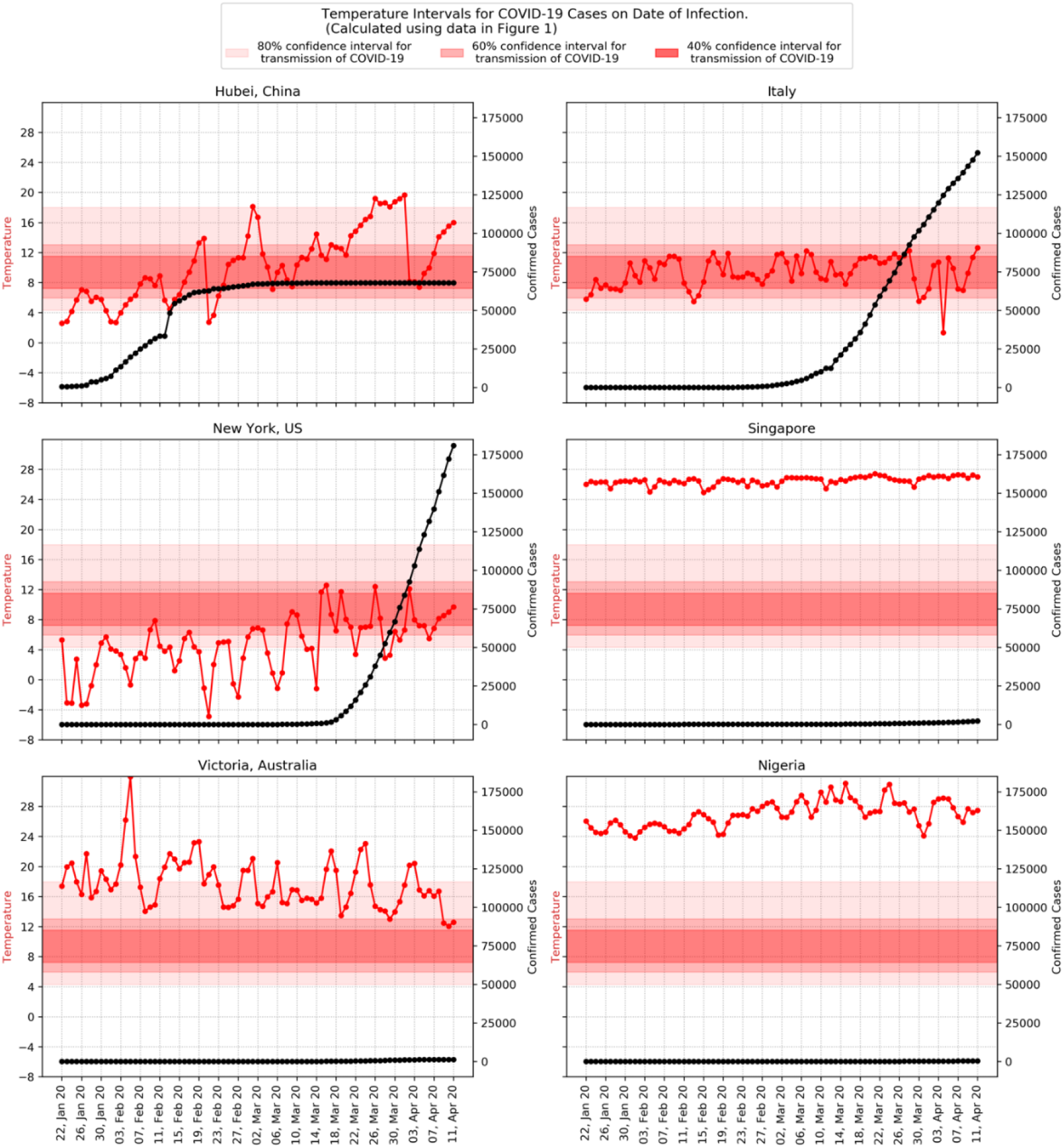
These graphs show changing temperatures on the y-axis (red line) for Hubei (China), Italy, New York, Singapore, Nigeria and Victoria (Australia) from 22^nd^ of January 2020 to 11^th^ April 2020 (x-axis). Also shown are the 40%, 60% and 80% CI for the median worldwide temperature for COVID-19 outbreaks (9.12°C). Superimposed on each graph is the total confirmed cases of COVID-19 (black line) at each location, quantitated on the y-axis on the right. These demonstrate the correlation between the regions with temperature ranges in the “red zone” and total number of cases.

## Discussion

Our analysis shows a clear association between temperature and enhanced risk of Sars-Cov-2 transmission with the majority of world cases and deaths occurring at temperatures between 4°C and 18°C. This temperature-outbreak relationship is consistent with other human coronaviruses (HCoV-229E, HCoV-HKU1, HCoV-NL63, and HCoV-OC43) which cause symptoms mainly in cooler winter periods and rarely occur during summer.^12–14^ Furthermore, SARS-CoV responsible for the 2003 SARS, also demonstrated enhanced transmission at cooler temperatures.^5–7^ Lower temperatures have been associated with seasonality of other viruses such as influenza and respiratory syncytial virus, which surge during colder winter periods.^15–18^ The reasons for this enhanced viral spread during colder weather may be due to enhanced shedding of the virus during colder weather^17,19^, stability of the virus suspended in air droplets^20^ or increased viability on nasal mucus membranes cooled by surrounding air.^21^ Additionally, cold weather can impair the immune system by inhibiting nasal mucociliary clearance and phagocytosis, diminishing the protective properties of mucus in upper airways^22^, or predispose to vitamin D deficiency due to lower exposure to ultraviolet radiation making people more susceptible to the infection.^23^

Based on our analysis, countries should incorporate ambient temperature forecasts into account when developing risk prediction models and even calculating R-nought (number of people one infected person subsequently infects). With no country yet reporting more than 1% of the population infected, the potential beneficial public health effects of “herd immunity” (at-risk people protected from infection because they are surrounded by immune individuals) is still out of reach and it is likely that it will be months to years before a vaccine becomes available for widespread use.^24^ Therefore, strict application of public health strategies such as lockdowns, social distancing, testing and contact tracing should be strongly considered for countries heading into temperatures that range between 4°C and 18°C. These measures should also be continued in countries currently experiencing temperatures in this range to assist the prevention of emergence/re-emergence of the epidemic. This is important, as a vast proportion of people infected with SARS-CoV-2 can remain asymptomatic and freely mix with the general population, transmitting the virus.^25–27^

Extrapolating the 2019 temperature data (as shown in Figure 3) for the remaining 2020 year, it is possible to predict when regions may approach this dangerous temperature range for enhanced viral transmission. For example, Victoria, Australia will enter this band in mid-May and remain in it until mid-October. Later in the year, with the onset of the Northern hemisphere winter, New York, Italy and Hubei will be in the band again from late October onwards. The identification of these at-risk time periods will aid governments and public health authorities in planning their surveillance and monitoring activities to counter the risk of recurrence of SARS-CoV-2 outbreaks

## Strengths and limitations

The key strength of our study is that we included the entire worldwide population of cases and deaths over a 3 month time period. This avoids sampling errors and makes the demonstrated association between temperature and COVID-19 outbreaks significant. Some COVID-19 cases may have travelled during the incubation period and were subsequently detected, recorded and attributed to another region with a different temperature. These people movements were not taken into account and may potentially add unintended bias to our findings. We have taken the temperature of each region at its respective capital, but there is likely to be some temperature variance among various locations around the capital region. This may serve to obscure correlations between temperature and COVID-19 cases and deaths. We chose to use the average daily temperatures for our analysis, but acknowledge that this may not represent the exact outdoor temperatures on particular days when people became infected. Studies showed that the median incubation period of SARSCoV-2 is 5 days^9^ and the mean time to death from symptom onset was 18 days^10^ and we used these numbers in our analysis. However, the incubation period can vary from 2–14 days with 1% of cases occurring after 14 days and time to death can vary according to the clinical course of the disease.^9^ This may introduce some imprecision in to the histogram that we presented, but as temperatures are unlikely to change significantly over the course of a few days, the overall impact of these variations is likely to be minimal. Some regions may have under-reported cases due to, for example, lack of available testing or being unaware of their asymptomatic carriers. This will likely underestimate the peak infection incidence as regions with the largest outbreaks will have the highest demand for tests or simply assume people are infected without confirmation testing. Our analysis also does not take into account public health measures such as mandatory lockdowns, social distancing and contact tracing measures that could minimize outbreaks.^28^ Other environmental factors such as humidity, windspeed and ultraviolet radiation levels as well as regional factors such as population density and social behaviour may also affect the transmission of coronaviruses and its relationship with ambient temperature.^5^

Whilst we are aware that some warmer countries such as Singapore have had clusters of outbreaks^29^, this may relate to foreign imports, spread in air-conditioned buildings maintained at lower temperatures or transmitted from person to person via a fomite harbouring in cooler temperatures such as in refrigerators.^30^

## Conclusion

Our analysis shows a definitive association between rates of infection and death from COVID-19 and ambient temperature, with the highest risk being around 9°C. We suggest that countries take ambient temperature trends into account when planning policy on containment strategies such as social distancing and lockdowns to minimize future outbreaks of COVID-19. Further studies to assess potential relationships between COVID-19 infection and other environmental factors such as humidity, wind speed and ultraviolet radiation levels may better prepare us to tackle our invisible enemy.

No funding was received for the preparation of this manuscript.

All authors have completed the ICMJE uniform disclosure form at www.icmje.org/coi_disclosure.pdf and declare: no support from any organisation for the submitted work; no financial relationships with any organisations that might have an interest in the submitted work in the previous three years; no other relationships or activities that could appear to have influenced the submitted work.

The Corresponding Author has the right to grant on behalf of all authors and does grant on behalf of all authors, a worldwide licence to the Publishers and its licensees in perpetuity, in all forms, formats and media (whether known now or created in the future), to i) publish, reproduce, distribute, display and store the Contribution, ii) translate the Contribution into other languages, create adaptations, reprints, include within collections and create summaries, extracts and/or, abstracts of the Contribution, iii) create any other derivative work(s) based on the Contribution, iv) to exploit all subsidiary rights in the Contribution, v) the inclusion of electronic links from the Contribution to third party material where-ever it may be located; and, vi) licence any third party to do any or all of the above.

Patients or the public were not involved in the design, or conduct, or reporting, or dissemination plans of our research.

## Data Availability

All data freely available to anyone at anytime at the links attached.

https://coronavirus.jhu.edu/map.html

## Summary boxes

### What is already known on this topic

The World Health Organization situation reports on COVID-19 show that regions which have experienced cooler temperatures such as Tehran, Milan and Madrid in February and New York and London in March have experienced the highest burden of disease.
Warmer regions in South East Asia, Africa and Australia have had the lowest outbreaks.
Whilst laboratory studies and epidemiological data on other coronaviruses have shown a link between cooler temperatures and higher rates of coronavirus transmission, a putative association between temperature and SARS-CoV-2 infection rates was yet to be convincingly established.

### What this study adds

Our results show a clear association between the rates of infection and death from COVID-19 and ambient temperature, with the highest risk occurring around 9°C.
These findings provide strong support for the role of ambient temperature in determining where and when significant outbreaks of COVID-19 infection occur.
This information may be vital for governments and public health authorities when determining their policies for containment, mitigation and surveillance of COVID-19 outbreaks; for example, a strategy to relax measures such as social distancing may be best avoided at times when ambient temperatures correspond to the highest risk of transmission.
Our results also encourage the utilization of temperature information as a critical component in current and future risk-prediction models.

## References

1. Zhu N, Zhang D, Wang W, et al. A Novel Coronavirus from Patients with Pneumonia in China, 2019. N Engl J Med 2020; 382(8): 727–33.

2. World Health Organization. Coronavirus disease 2019 (COVID-19) Situation Report –83. 2020; Available at: https://www.who.int/docs/default-source/coronaviruse/situation-reports/20200412-sitrep-83-covid-19.pdf?sfvrsn=697ce98d_4. Accessed April 13th, 2020.

3. Casanova LM, Jeon S, Rutala WA, Weber DJ, Sobsey MD. Effects of air temperature and relative humidity on coronavirus survival on surfaces. Appl Environ Microbiol 2010; 76(9): 2712–7.

4. Chan KH, Peiris JS, Lam SY, Poon LL, Yuen KY, Seto WH. The Effects of Temperature and Relative Humidity on the Viability of the SARS Coronavirus. Adv Virol 2011; 2011: 734690.

5. Cai QC, Lu J, Xu QF, et al. Influence of meteorological factors and air pollution on the outbreak of severe acute respiratory syndrome. Public Health 2007; 121(4): 258–65.

6. Lin K, Yee-Tak Fong D, Zhu B, Karlberg J. Environmental factors on the SARS epidemic: air temperature, passage of time and multiplicative effect of hospital infection. Epidemiol Infect 2006; 134(2): 223–30.

7. Tan J, Mu L, Huang J, Yu S, Chen B, Yin J. An initial investigation of the association between the SARS outbreak and weather: with the view of the environmental temperature and its variation. J Epidemiol Community Health 2005; 59(3): 186–92.

8. Yao Y, Pan J, Liu Z, Meng X, Wang W, Kan H, Wang W. No Association of COVID-19 transmission with temperature or UV radiation in Chinese cities. Eur Respir J 2020.

9. Lauer SA, Grantz KH, Bi Q, et al. The Incubation Period of Coronavirus Disease 2019 (COVID-19) From Publicly Reported Confirmed Cases: Estimation and Application. Ann Intern Med 2020.

10. Verity R, Okell LC, Dorigatti I, et al. Estimates of the severity of coronavirus disease 2019: a model-based analysis. Lancet Infect Dis 2020.

11. United Nations, Department of Economic and Social Affairs, Population Division (2019). World Population Prospects 2019: Volume I: Comprehensive Tables.

12. Gaunt ER, Hardie A, Claas EC, Simmonds P, Templeton KE. Epidemiology and clinical presentations of the four human coronaviruses 229E, HKU1, NL63, and OC43 detected over 3 years using a novel multiplex real-time PCR method. J Clin Microbiol 2010; 48(8): 2940–7.

13. Dowell SF, Ho MS. Seasonality of infectious diseases and severe acute respiratory syndrome-what we don’t know can hurt us. Lancet Infect Dis 2004; 4(11): 704–8.

14. Hendley JO, Fishburne HB, Gwaltney JM. Coronavirus infections in working adults. Eight-year study with 229 E and OC 43. Am Rev Respir Dis 1972; 105(5): 805–11.

15. Chew FT, Doraisingham S, Ling AE, Kumarasinghe G, Lee BW. Seasonal trends of viral respiratory tract infections in the tropics. Epidemiol Infect 1998; 121(1): 121–8.

16. Tamerius JD, Shaman J, Alonso WJ, Bloom-Feshbach, K, Uejio CK, Comrie A, Viboud C. Environmental predictors of seasonal influenza epidemics across temperate and tropical climates. PLoS Pathog 2013; 9(3): e1003194.

17. Lowen AC, Mubareka S, Steel J, Palese P. Influenza virus transmission is dependent on relative humidity and temperature. PLoS Pathog 2007; 3(10): 1470–6.

18. Jaakkola K, Saukkoriipi A, Jokelainen J, et al. Decline in temperature and humidity increases the occurrence of influenza in cold climate. Environ Health 2014; 13(1): 22–069X.

19. Pica N, Chou YY, Bouvier NM, Palese P. Transmission of influenza B viruses in the guinea pig. J Virol 2012; 86(8): 4279–87.

20. Jaakkola K, Saukkoriipi A, Jokelainen J, et al. Decline in temperature and humidity increases the occurrence of influenza in cold climate. Environ Health 2014; 13(1): 22–069X.

21. Pica N, Chou YY, Bouvier NM, Palese P. Transmission of influenza B viruses in the guinea pig. J Virol 2012; 86(8): 4279–87.

22. Lowen AC, Steel J. Roles of humidity and temperature in shaping influenza seasonality. J Virol 2014; 88(14): 7692–5.

23. Zittermann A, Pilz S, Hoffmann H, MÃ¤rz, W. Vitamin D and airway infections: a European perspective. Eur J Med Res 2016; 21: 14–016.

24. Amanat F, Krammer F. SARS-CoV-2 Vaccines: Status Report. Immunity 2020; 52(4): 583–9.

25. Gudbjartsson DF, Helgason A, Jonsson H, et al. Spread of SARS-CoV-2 in the Icelandic Population. N Engl J Med 2020.

26. Sutton D, Fuchs K, D’Alton, M, Goffman D. Universal Screening for SARS-CoV-2 in Women Admitted for Delivery. N Engl J Med 2020.

27. Mizumoto K, Kagaya K, Zarebski A, Chowell G. Estimating the asymptomatic proportion of coronavirus disease 2019 (COVID-19) cases on board the Diamond Princess cruise ship, Yokohama, Japan, 2020. Euro Surveill 2020; 25(10): 2000180. doi:10.2807/1560.

28. Prem K, Liu Y, Russell TW, et al. The effect of control strategies to reduce social mixing on outcomes of the COVID-19 epidemic in Wuhan, China: a modelling study. Lancet Public Health 2020.

29. Pung R, Chiew CJ, Young BE, et al. Investigation of three clusters of COVID-19 in Singapore: implications for surveillance and response measures. Lancet 2020; 395(10229): 1039–46.

30. Bedford J, Enria D, Giesecke J, et al. COVID-19: towards controlling of a pandemic. Lancet 2020; 395(10229): 1015–8.

